# The complexities and contradictions of Online Eating Disorder Content: A qualitative evidence synthesis of young people’s views and experiences

**DOI:** 10.1101/2025.10.12.25337834

**Authors:** Meena Khatwa, Rebecca Rees, Claire Stansfield, Vidhi Bassi, Ceri-Nicole Wilkins, Kelly Dickson

## Abstract

Eating disorders (ED) continue to be a concern worldwide. Traditional media is seen as contributing to problematic discourses about ideal bodies. However, more recently, concern has grown about content and spaces that are online, and if they increase disordered eating and other ED symptoms.

A qualitative evidence synthesis was conducted to explore young people’s lived experiences of online ED content. Studies were included if they provided findings about views that related online content to body image concerns or eating disorder symptoms from young people from OECD countries aged 11-25 years old (from 2007 onwards).

This article focuses on findings from eight included studies whereby three key themes were identified: Comparing - how online content was used to evaluate others and oneself; Curating - how young people manage this content; and Community - spaces for shared experience and aspirations.

Young people illustrated the complex ‘double-edged’ nature of online spaces, which can offer inclusion but can also be experienced as harmful. The synthesis outlined a lack of supportive offline mental health-promoting spaces as part of the context of young people’s online content use.

## INTRODUCTION

The rise of online eating disorder content (OEDC) is a growing concern due to the increased use of social media platforms and online communities (Saul et al. 2022). To understand the complexities and contradictions of OEDC and its relationship to body image we have considered eating disorders (ED), such as anorexia nervosa (AN), bulimia nervosa (BN), and binge eating disorder (BED), and how they present in different ways but are all characterised by distress or concern about body image and eating (1). Though eating disorder symptoms can be detected from as young as six into late adulthood; peak onset predominately occurs in early adolescence as does increased use of social media and engagement in online settings (2, 3, 4).

Higher incidence of eating disorders is reported in girls and young women, however, emerging research with boys and young men has begun to identify specific disordered eating and weight gain behaviours related to body ideals that emphasise muscularity and leanness, and which can lead to elevated ED symptoms (5, 6). Globally, it is estimated that over 3.3 million healthy lives are lost due to eating disorders, with increases in both the risk of suicide and premature death, with the highest mortality rates occurring in young people with anorexia (7, 8, 9). Living with an eating disorder can be highly disruptive, exerting a negative impact on a young person’s sense of self, relationship with others and overall quality of life (10, 11). Similar concerns have also been raised in relation to the amount of time children and young people spend in online spaces (Dickson et al. 2019).

The factors contributing to an eating disorder in young people are caused by a complex interaction of bio-psychosocial and environmental mechanisms (12, 13). A key area of investigation has evolved to explore the juxtaposition between how sociocultural views about ‘ideal’ bodies interact with genetic and other factors (for example, personality traits, family environment, genetics) to increase the risk of an eating disorder (14, 15). Historically, traditional media (for example, television, films, magazines) have been theorised as a social arena that generate and reproduce discourses about ideal body images (16). This has led to a large volume of empirical research on exploring the effects of mass media on eating disorders in children and young people (17, 18). In the advent of the internet, this literature has now extended to consider the impact of digital media on young people’s body dissatisfaction and disordered eating (19)

A distinctive feature of online digital media platforms (e.g., websites, online forums, and social networking sites), is their interactivity. They enable young people to be generators and receivers of personal and targeted content, often simultaneously (20). A recent trend in this direction has been the development of both online pro-eating disorder and eating disorder recovery content. Whilst pro-ED content actively encourages disordered eating behaviours, pro-recovery content aims to support and promote getting better (21, 22). With the increase in new types of digital media, there have been growing concerns about the ‘widespread existence of pro-ED communities (23) and their accessibility and expanding scope of ED content, which, no longer confined to websites and online forums. This content is being produced and reproduced in more sophisticated ways via digital media platforms (for example, Instagram, Tumblr, YouTube, Pinterest, Facebook and Tiktok) (22, 24, 25, 26).

Considering the complexity, this qualitative evidence synthesis adopted a broad scope driven by young people’s own definitions. Thus, online eating disorder content (OEDC) was defined as any material or exchanges identified by young people as holding the potential to influence their own or others’ distress or concern about body image or about behaviours related to body size, shape, or weight. This supported us to answer the following research question: What are young people’s views and experiences of online eating disorder content, and what influence does it have on their relationship with their bodies, weight control behaviours, and/or any other eating disorder symptoms?

## METHODS

A review of research was undertaken which included a qualitative evidence synthesis (QES). The protocol was published on the <u>PROSPERO</u> systematic review registry [anonymity no author]. The review work was informed by two consultative workshops with a group of young people with experience of accessing OEDC. The methods used for these workshops and all aspects of the review, are detailed fully in a technical report.

### Search strategy

#### Eligibility criteria and study selection

For studies to be included in the QES, they needed to use qualitative methods to investigate young people’s experience and views of online eating disorder or body-image content. The sample needed to be aged between 11-25 and identify as having an eating disorder. Studies also needed to be conducted in an OECD country and published in English from 2007 onwards. A comprehensive search was undertaken to comprehensively identify research on body dissatisfaction, disordered eating or over-exercising symptoms and behaviours where it focused on online content, social media, blogs or instant chat involving young people. This involved searches of 15 scholarly bibliographic databases in health, psychology and social science, eight online resources and search engines, four online journals, and forward and backward citation searching. Two update searches were run (the last in October 2024) to seek additional, more recent studies.

Search results were imported into the review software, EPPI-Reviewer 4 (27), which was also used to manage all subsequent stages of the review described below. References were de-duplicated, then screened initially on titles and abstracts. Full reports were obtained for those references judged as meeting the inclusion criteria or where there was insufficient information from the title and abstract to assess relevance. A second opinion was made available for any study where a reviewer was unsure of its eligibility. We piloted the exclusion criteria by comparing decisions in groups of two or more reviewers using worksheets with guidance notes on a small sample of records (for example, 10-20).

Disagreements were resolved through consensus and any required refinements to the criteria were made and recorded in a working protocol document. A further sample of records were screened by reviewers independently and differences resolved by discussion or consulting with a third reviewer. When agreement was adequate (90-95%) for this second sample, the remaining citations were screened by a single reviewer.

#### Data Extraction

The coding tool used for extraction were devised from previous reviews (Dickson et al., 2018, Lester et al., 2019). Codes captured geographical location, population, study design. study aims, and sampling and data collection methods, types of online content, platform, and ED, as well as an extraction of the authors’ summary description of their findings. Two reviewers [MK & RR] independently piloted the second tool. Each study was then coded independently by two reviewers who then reached consensus to produce a final agreed coding.

#### Quality assessment of primary studies

The appraisal was of the primary studies was done independently by two of the authors, who then reached a consensus. The allocated studies were critically appraised against six sets of questions used in previous QES reviews (e.g. anonymity no author] These questions explored each study’s approach to: (1) sampling; (2) data collection; and(3) data analysis; as well as (4) the study’s grounding of findings in young people’s perspectives; and (5) the breadth and depth of the findings. Study quality was summed up in terms of: reliability (which considered the judgements made using criteria 1-4; and usefulness (which used questions 5 and6).

#### Data synthesis

Data was synthesised thematically (28), and included qualitative data consisting of participants quotes and authors interpretation direct from primary studies. The process involved line-by-line coding by two reviewers [MK & RR] and was carried out independently using the coding function in EPPI-Reviewer (web-based software programme). By using an iterative process, the coding of the data was then organised into themes whereby higher-order themes were identified through repeated individual coding and follow-up discussions between these two reviewers. The discussions between the reviewers led to the synthesis being divided into two parts due to the complexity and breadth of the data. The team first worked on coding and drafting a narrative synthesis for 13 studies of views from young people with lived experience of EDs. Two of the young people who had participated in the workshop [BV & CNW] also worked on this synthesis to help incorporate the findings of the four most recently published studies. A second synthesis (not reported here) was conducted on studies where young people didn’t have lived experience.

## RESULTS

After the removal of duplicates a total of 10484 references were screened on title and abstract and 693 references were rescreened on a basis of the full-text reports. Figure 1 illustrates the review’s screening process. A total of 13 studies, reported in 19 papers were included in the review. See Table 1 for an overview of these studies. Supplementary Files A and B provide additional detail.

**Figure 1:** Flow of literature through the review.

**Table 1.** Overview of studies characteristics included in the review.

| Author | Study population<br>(location/sample size; ED-related characteristics)* | Type of content and online platform type under discussion |
| --- | --- | --- |
| **Dashow, 2021** | •USA; n=12<br>•Had ED recovery focused accounts | •Online Content focus: Recovery<br>•Online platform: Instagram |
| **Eikey & Booth, 2017** | •USA; n=16<br>•Formally diagnosed with ED and/or have had symptoms of anorexia or bulimia. Most reported being in recovery | •Online Content focus: Body Image Ideals, Fit, Thin<br>•Online platform: Websites, Instagram |
| **Faccio, 2024** | •Italy; n=30<br>•With a diagnosis of an ED, and in the early stages of hospital treatment | •Online Content focus: Body Image Ideals, Self-representation<br>•Online platform: Instagram and Snapchat |
| **Firkins, 2017** | •UK & USA; n=3<br>•A history of eating difficulties and previous regular use of pro-ED sites now reduced or stopped altogether | •Online Content focus: Pro-eating disorder<br>•Online platform: Websites, social media |
| **Hockin-Boyers et al., 2021** | •UK; n=19<br>•A history of eating disorders, now weightlifting during recovery | •Online Content focus: Body Image Ideals, Fit (transformation photos)<br>•Online platform: Instagram |
| **IPSOS, 2024** | •UK; n=8***<br>•Self-identifying with an ED, with at least six months since discharge from an ED support service and without an active ED diagnosis | •Online Content focus: Content perceived as harmful by participants<br>•Online platform: Not pre-specified |
| **Mincey, 2019** | •USA; n=10<br>•Accessing and using pro-eating disorder online media at the time of the study | •Online Content focus: Pro-eating disorder<br>•Online platform: Websites, social media (forums) |
| **Nikolova & LaMarre, 2023** | •New Zealand; n=8<br>•In recovery/on a journey to recovery from disordered eating and using Instagram 3+ times a week | •Online Content focus: Health and fitness<br>•Online platform: Instagram |
| **Praet, 2024** | •Belgium; n=7<br>•With a DSM-5 diagnosis of Anorexia, restrictive type, recruited through a paediatrics department | •Online Content focus: Body Image Ideals, Pro-eating disorder, Recovery<br>•Online platform: Social media |
| **Saunders et al., 2020** | •USA; n=15<br>•Self-identifying as in ED recovery | •Online Content focus: Body Image Ideals: Beauty/Attractiveness (selfies)<br>•Online platform: Social media |
| **Sjöström, 2024** | •Sweden; n=14<br>•With a DSM-5 diagnosis of Anorexia, recruited through an ED clinic. | •Online Content focus: Recovery<br>•Online platform: TikTok |
| **Smahelova et al., 2020** | •Czech Republic; n=30<br>•Experience (current or past) of an ED | •Online Content focus: No restriction<br>•Online platform: Social media, Websites |
| **Yeshua-Katz and Martins, 2013** | •USA, UK, Australia, New Zealand, the European Union; n=33<br>•Running a Pro-anorexia blog, small number reported being in recovery, most reported living with the ED | •Online Content focus: Pro-anorexia<br>•Online platform: Social media (blogs) |

\* With the exception of one young man in Mincey (2019) and an unspecified number in IPSOS and Tonic (2024), all participants were female. \*\* Some of the findings in IPSOS and Tonic (2024) could not be used for our review as participants included young people who did not necessarily have previous lived experience of eating disorders. Some were described as having experienced self-harm, suicidal ideation, anxiety and/or depression, or as having encountered content that they perceived as harmful for these conditions.

### Design of included studies

The 13 studies had all been published since 2017, indicating this is a relatively new field of qualitative enquiry. A range of OECD countries were focused upon, including the USA (4 studies), the UK (2 studies), and Belgium, the Czech Republic, Italy, New Zealand and Sweden (1 study each). Two additional studies recruited participants from across several countries.

In terms of the kinds of young people participating, only one study reported clearly that it involved young men with lived experience of EDs, and only one participant in this study fell within this review’s age range focus (29). A range of ages were represented, with seven studies’ samples including participants aged less than 18 (30, 31, 32, 33, 34, 35, 36). Although studies collected information about class and ethnicity, exploration of disadvantage, such as how ethnicity or socioeconomic status might intersect with subjective experiences of engaging with online content, was not a focus in any of the studies. In terms of content, four of the 13 studies explored pro-ED focused content, or content perceived as “harmful”, one focused upon recovery-focused content, while the remainder focused on more general content promoting fitness, health and/or body-image ideals, or were open to discussion of any of these content types. In terms of online platform types, one study focused upon TikTok and four on Instagram (one also exploring Snapchat), while others focused upon blogs, the use of selfies and social media more generally.

### Quality of studies and consultation with young people

The quality of studies is summarised in Table 2. All but one study was judged to be of at least medium reliability and/or usefulness for the purposes of this qualitative evidence synthesis.

**Table 2.** Quality assessment of studies: Reliability and usefulness.

| Author | Reliability |  |  | Usefulness |  |  |
| --- | --- | --- | --- | --- | --- | --- |
|  | High | Medium | Low | High | Medium | Low |
| Dashow 2021 |  | ✓ |  |  | ✓ |  |
| Eikey & Booth 2017 |  |  | ✓ |  |  | ✓ |
| Faccio et al., 2024 |  | ✓ |  |  | ✓ |  |
| Firkins et al., 2017 |  | ✓ |  | ✓ |  |  |
| Hockin-Boyers et al., 2021 | ✓ |  |  | ✓ |  |  |
| IPSOS 2024 |  | ✓ |  |  |  | ✓ |
| Mincey, 2019 |  | ✓ |  |  | ✓ |  |
| Nikolova & LaMarre, 2023 |  | ✓ |  |  | ✓ |  |
| Praet et al., 2024 |  | ✓ |  |  | ✓ |  |
| Saunders et al., 2020 |  | ✓ |  |  | ✓ |  |
| Sjöström et al., 2024 | ✓ |  |  | ✓ |  |  |
| Smahelova et., Al 2020 | ✓ |  |  | ✓ |  |  |
| Yeshua-Katz & Martins 2013 |  | ✓ |  | ✓ |  |  |

In a first workshop before searches were finalised, there was a discussion of the kinds of OEDC and language that young people might be using and what mattered most for young people about OEDC. The young people discussed emerging findings from the synthesis at the second workshop. They identified aspects that they considered important but that might have been missed by the research team when coding the included studies. These included OEDC use for self-validation, and sites that present as focused on ED recovery. The reviewers added detail about both to the synthesis, aided by the addition of new studies found after update searches were run.

### Thematic Synthesis Findings

There were three distinct overarching themes developed from carrying out the qualitative synthesis: (1) Comparing, which illustrated how young people use content to evaluate their own and others’ bodies and behaviours. (2) Curating, which explored how young people manage and create content, and (3) Community, which examined the reasons why young people were drawn to social spaces where content is shared, and what purposes these serve.

### Comparing

All 13 studies explored how young people used ED content found on diverse online platforms to evaluate others and themselves (29, 31, 32, 33, 34, 35, 36, 37, 38, 39, 40, 41, 42). This theme and the sub sections that follow cover a range of complex behaviours and experiences which include types of content used to compare such as weight loss and body image; how content which is easily accessible can trigger negative thoughts such as failure but also feelings of affirmation. It is also pertinent to highlight that young people’s experiences were influenced by where they were in their eating disorder journey, and accounts of online comparisons in recovery were sometimes positive, but not always.

#### Comparisons are constant – “seeing thin people on my feed helps” (37)

There were many types of content posted online which illustrated a comparison context. This included: Calorie counts or the size of food portions, accounts of exercise taken (36, 37), popularity which resulted in the form of views or likes (Saunders et al., 2020), as well as images of bodies (31, 36, 37, 38, 43). This content could be found on platforms which included anorexia-focused and pro-recovery forums (29, 31, 37, 38) but also on social networking platforms including Instagram (e.g. Eikey), Snapchat (e.g. (33) and TikTok (36) and other social networking sites (35, 36). The 13 studies also revealed that ease of access to undesirable online content could lead to feelings of consuming too much. Young people used terms like “constantly” (30, 37), “bombarding” and “attacking” (44) and “[getting] hooked” (36). when describing this experience. One study’s authors (42) described how users in recovery were critical of algorithms within Instagram which could ‘misinterpret’ what young people wanted to see, treating diet content and body-positivity as synonymous (e.g. sending diet-recommendations as opposed to encouragement not to moralise around food). These experiences coupled with the ubiquitous nature of comparative online words and images meant they were also implicated as potential triggers of negative emotions (29, 30, 31, 32, 33, 35, 36, 41, 42). This was particularly troubling for some young people in their recovery or treatment phase of the ED journey, whereby comparing to others led to weight loss behaviours and “pulled them back to the illness” (31). Relatedly, bloggers (32) reported feeling responsible for potentially triggering others (32). This was juxtaposed with others actively seeking such triggering material online, so as to “actively engage in ED behaviour” (29, 45), for example, focusing repeatedly on the food in videos to make it easier not to eat (36).

#### Comparison can affirm but also undermine sense of self – “this person is skinnier than me so she’s sicker than me” (29)

Viewing and interacting with online content could also conjure up complex feelings which could shift between personal affirmation and failure. There was reference to how seeing others’ content could strengthen existing feelings of not being sick enough to deserve support (e.g. see quote in this section’s header). Online comparisons could influence and lead to feelings of low self-esteem whereby, when viewing others’ accounts of their behaviours or bodies, participants felt triggered (34, 35) or like “losers”, in that “they are doing a better ‘job’ of having an eating disorder” (29) The potential for “negative inspiration” was also emphasised, where “pro-ana blogs represented EDs as a desirable lifestyle” (44). However, the emotional impact of looking at online content when comparing could also be positive, when in recovery. Young women reported seeing online body transformation, where others went from underweight to deliberately gaining muscular weight, or ‘becoming body builders’, and accounts documenting self-transformation were celebrated (37). (39) It was striking that the feelings expressed about comparisons by young people very much depended upon where they were in their eating disorder journey, being generally more positive for those identifying as in recovery (29, 30, 43, 44).

### Curating

This overarching theme captures different levels of engagement which go beyond merely observing and include the management and creation of content. The theme considers activities such as: use of hashtags or other means for seeking out certain content producers or types of content; liking and providing feedback to others’ individual posts; and otherwise interacting with existing content. Young people in the studies also deliberately created new material, such as uploading images on Instagram (annotated or otherwise), running Instagram recovery accounts, posting questions and responses in specialised forums, and writing blog articles. The following sub-themes highlight the key findings about, in turn, what is interpreted as curation, what young people engage with and why and how this impacts behaviour and lived experiences of ED.

#### Seeking out and creating content that is authentic – “I think it gives us more access to real people” (30)

One element of curation that resonated with young people was the seeking out of online ED content that they could relate to alongside their own journeys. Young people searching for pro-ED material expressed a desire for following “concrete – real people with ED and their stories” (31). In part this aided participants’ motivation for recovery because they had realistic examples “where real people [on social media…] are having the same thoughts” (30) rather than in the past where most content that was found in magazines was of celebrities only. The value of authenticity is also shared by blog creators who in one study, were keen to: “share and declassify,” or to open up about themselves (44). This sharing included young people reporting on their whole life and not just their ED behaviour; they positioned their posts “within their everyday life and family context” (31). The complexity of this act is indicated by one young woman, who shared: “I often refer to how my blog is my true self and how in real life I have to act and hide my true feelings.[…]. My blog is who I am backstage when I’m stripped of the make-up and costume” (32).

Positive experiences of authentic content were countered by views of negative impacts associated with inauthentic content, such as image manipulation (35). The unhelpfully selective sharing of content was implicated in concerns about “people lying to themselves … and lying to others that they were recovering” (44, 46).

#### Managing and interacting with others’ content – “it’s not a good or a bad thing, it’s a tool” (30)

Young people described a wide range of topics covered when responding to others’ content. They also commented upon the intensity of interactions and the amount of time spent in exchanges, which could both be high (31, 41). A response to an individual’s post could be seen as a signal of support by young people, whether this was in pro-ED discussion sites (29, 31) or in recovery-focused discussions (37, 39, 41, 43): “If I have a friend who lost a lot of weight. I’ll always comment … “you’re on a good path, you’ve done this in a healthy way” “(37). However, some responses aimed to encourage self-reflection. for example, in response to transformation photos on Instagram, one participant in recovery recalled posting how weight loss didn’t make a person happier (39). Some young people were selective with who they followed and unfollowed (30, 36) when content was considered unhelpful or, as one young woman put it, when “what you’re posting isn’t contributing positively towards my mental health” (42). All participants in one study reported unintentionally encountering algorithmically recommended harmful content (34). They also reported being wary when seeing extremely low calorie diets or fasting, but found it difficult to distinguish more generally between healthy lifestyle and pro-ED content. Others were careful with the platform’s like function (36). However, young people also identified reduced control over triggering content, reporting feelings of addiction (35), as well as algorithmic content coming “from nowhere”, and requirements for “endless scrolling” (36). Others found it difficult to “filter out” things they didn’t want to see or disconnect from friends who demonstrated “toxic behaviour” (42).

### Varied motivations for generating new content – “it’s like to say it out loud” (37)

The studies revealed there were many motivations/factors for creating new content across different platforms (content other than annotations or direct responses to others’ material). For instance, bloggers who were living with ED placed an emphasis on producing content as a means of self-expression, gaining support (not feeling alone), coping with stigma and raising awareness of shared lived experiences (31, 32, 37, 41). Online spaces allowed young people to share their own ED stories with less danger of being judged: as one young woman put it “having an ED was so socially unacceptable that it pushed me to seek others that I could tell my story to” (32). Describing the therapeutic properties of disclosure, one young woman noted, “there really happened an extreme disclosure and I think it was one of the most important steps (for recovery) … by putting it down in written form, and actually identifying with it … It’s like to say it out loud (31) Writing and sharing recovery content not only helped the bloggers but was seen as helping others. As well as self-expression through written content, young people also described the sharing of photos of visually appealing healthy meals or body transformations. (39). For example, young people who were weightlifting as part of their recovery reported comparisons between their own before and after selfie photos to be helpful, one commenting, “when I look back at what I used to look like and what I look like now it helps me be, like, ok. I am going in the right direction and other people can see it” (31, 39). Some pro-ED bloggers also reported writing to counteract inaccurate portrayals of EDs, to “make them realize that it’s not what everybody makes it out to be” (32).

### Community

The findings in this theme provide insight into why young people are drawn to online virtual spaces, whereby a sense of community connections is evoked through shared identification, experiences, and aspirations. However, these ED communities also appear to be complex spaces, which can be both helpful and harmful, thus creating a double-edged sword. Again, it was evident that a young person’s level of engagement was influenced by their ED journey. For instance, in the early stages an individual might be curious or seeking connections. Or perhaps further along the way there could be impacts on physical and emotional wellbeing, and disengaging could be complicated.

### Combatting loneliness and being misunderstood – “nobody normal understands why you want to starve yourself” (32)

The attraction of online communities was referred to in ten of the 13 studies, where young people with experience of EDs revealed feelings of isolation, loneliness and being misunderstood by the offline world (29, 30, 31, 32, 36, 38, 41, 42). In contrast, these young people described how the online world offered spaces that were less judgmental with the potential for both instructive and emotional support, and meaningful connection “with other people, like-minded people, so a sense of community … Others who have had a similar experience to me, and whose viewpoint or worldview aligns with mine” (42).

It seemed that young people felt there was no alternative for them when in a disordered state, and that the offline world did not offer what they needed (29, 31, 32, 36, 38, 41). As one young person put it, “I almost felt that I was an alien that I just didn’t fit in anywhere… I think it’s where I belonged and that’s where I felt comfortable… I didn’t feel like I was in turmoil there” (38). Another appeal of online communities was anonymity, this was described as a key driver of engagement where there was “no visual attachments” and therefore “easier to converse without feeling threatened” (32). However, anonymity was also mentioned as a way that people offering help could “degrade” others (34) and was thought to help “radicalize some people’s opinions” (44).

Some studies also highlighted limitations to acceptance within ED-related communities. For example, one young woman expressed that it was “not a healthy acceptance” when she had been in a disordered state. One young person, reflecting upon being part of a fitness recovery community, reported, “I don’t want to be, you know, the ‘eating disorder girl’ “ (39). Elsewhere, young people in recovery who reflected back described that the pro-ED platforms ‘could keep them in isolation’. In this study, all but one of the participants who had used pro-ED sites reported no longer using them (31)

### Sourcing support strategies – “I was only interested in how to endure so I did not have to eat” (31)

This quote and others across the studies illustrate how young people turned to online communities for information and learning. Pro-eating disorder sites were viewed as useful resources where practical tips and tricks and knowledge were shared by more experienced users, such as how to hide ED symptoms, and suppress appetites, and otherwise bring about weight loss (29, 31, 32, 35, 38). There was also reference to advice about avoiding “suffering from the condition” (29, 31, 32, 38). In one study, participants characterised this form of advice as harm reduction, and provided illustrations, for example: “Things like, after purging not brushing your teeth because you would brush the stomach acid around your mouth, and you’d rot your teeth faster. Things like that have been really helpful and probably why I still have all of my teeth.” (29, 31, 32, 38).

Young people described other kinds of learning from online ED content, (29, 31, 32, 38, 42). Specifically about the development of eating disorders (29), how to talk with others about EDs (42), self-acceptance and taking a healthy approach (36), as well as the recovery process (37). Recovery testimonials were seen as having potential to support transformation (35, 36).

Despite use of the term community, discussions of support and learning in all but two cases made no reference to anything other than individual actions. Young people with Instagram recovery accounts said they helped hold them accountable but said no more (46). Elsewhere, one participant referred to mentoring others and having been awarded by her peers the status of “advanced warrior… in recognition of her Pro-Ana successes” (38). In another study there was reference to the role of a moderator who maintains pro ED webpages, and how so-called Ana Coaches “would be called out in the thread”, presumably as a means of alerting others to danger (29).

### Online communities are a double-edged sword – “a thin line between receiving support and being triggered” (36)

This final sub-theme describes different initiations, and degrees of interaction within the online communities and how these intersect with the young person’s ED journey, emotions and behaviour. To add to the positive potential of avoiding isolation and learning for recovery described above, young people identify how online groupings can have negative impacts and how, at times, their online and offline journeys are nonlinear. In terms of journeys, young people described different experiences of seeking out pro-ED content and making connections (e.g. (34). This could begin with “lurking” or being “curious” (29). Others described primarily “looking for healthier recipes” (38), or how pro-ED content “popped up when … looking for other weight-loss things” (29). Young people also talked about the influence of ED content on the pathway towards a greater eating disorder. People posting content were also initially found and then followed through keyword searches (34) Following a YouTube channel enabled one young person’s eating disorder to “bloom” (38). One author described participants as having gone “deeper into illness … thanks to the anonymity, the presentation of extreme cases, and the safe environment where visitors are supportive and nice to each other” (31). Later on in their ED journeys, half of those with Instagram recovery accounts in one study felt they had been problematic for their own recovery and several noted that the pro-ana community was present even within the recovery community (46). In another study one young person described engaging with personal recovery accounts as ‘very unhelpful’, and her having to ‘stay clear of them since realizing at the very start … this is not good on any level’ (42) Experiences were not linear. One young woman described how her use would shift back and forward in a cycle between intense and frequent visits for ideas and support and stepping away to avoid “listen[ing] to everyone else’s problems” (38). This was particularly pertinent for people in recovery who still reported using pro-ana sites in various ways, such as revisiting to look for friendship (38).

## DISCUSSION

### Summary of findings and cross cutting themes

The main objectives were to collect, analyse and present evidence on young people’s experiences and views of engagement with eating disorder-related material on online platforms. As the research question proposed, the aim was to explore what kind of influences online eating disorder content has on young people’s relationships with their body, weight control behaviours, and/or any other eating disorder symptoms.

This synthesis of 13 studies of participants with direct experience of an eating disorder identifies diverse practices of comparing and curating online, and an emphasis on the value and challenges of being part of online ED-related communities. It starts to unpack the potential impacts online ED content can have on these young people. Recent quantitative primary studies have shown that social media platforms can be spaces of self-compassion and empowerment (47) but have also seen participants reporting the disadvantages and advantages of social networking (48).

Reduced levels of interaction with the offline world and a lack of external support/provision seemed to be a central experience of these young people. Online communities hold potential to offer refuge from the stigma and judgement of the offline world and to fulfil a need to belong and be understood. A 2015 study using a netnographic approach to explore posts within youth moderated online ED forums also echoed this notion in its identification of a “trusted environment for individuals to talk about isolation, fear… mutual support and acceptance” (49).

The use of pro-ana blogs was seen to provide emotional support, as other studies of online posts have shown (50, 51). Young people described embracing spaces where they could learn, comfortably self-evaluate, and share insights with others as to their shifting ED identities and therefore strengthen bonds (24). However, this is also juxtaposed with young people’s awareness of associated dangers. Pro-ED communities had been experienced as helpful but also damaging by young people.

The potential for online content to trigger unwanted behaviours seemed particularly pertinent in this group. Other studies have also found similar findings on triggering content (for example, (52). A level of awareness of potential triggers when producing and uploading content is also referred to by bloggers in one of the reviewed studies. This suggests a degree of responsibility and a perspective among online content producers that would benefit from further investigation. This review’s findings, however, should already illustrate for practitioners and policymakers how young people are active agents and producers of such content and not just passive consumers.

The findings from the QES also illustrate the contradictory and very complex nature of online experiences, thus evoking the idea of these environments comprising a ‘double-edged sword’ (53, 54). This phrase is also used by author (55) who discusses it in relation to “helping or harming one’s mental health and tendency towards eating disorders”. The shifting impact of eating disorder content across different stages of the disorder adds yet another layer of complexity. An understanding of this complexity could inform the work of social media providers in developing more effective detection and reporting tools for OEDC on their platforms, as well as educational materials for users. The studies also emphasised the importance of authenticity and how material might present distortions of reality. The sense of endless opportunities for body image -related comparisons and negative consequences suggests considerable commonality of experience for young people as a whole. The salience of community in young people’s engagement with online content could be incorporated into existing conceptual frameworks to enhance understanding of fluid and fluctuating engagement with content (56, 57). The observation of content, and the curating acts of following, commenting, and contributing content, are all likely to have different meanings for young people when done in the context of entering, engaging within, and leaving virtual social spaces where shared identity seems to be so key.

### Limitations of current research and future research

This review has revealed multiple terms used in the literature to describe the many complexities of eating disorders and body image. As reviewers, we have attempted to represent views and experiences by adhering to the ways that these have been expressed and reported in the included studies. For example, terms such as ‘Recovery’ were identified as a recurring theme as a main topic focus or what was shared by the young people. However, there was very little discussion by the young people studied of the use of recovery sites, or in-depth discussion of how different content and sites could impact on recovery. This could be considered limitations to the review, however, all reviews are necessarily limited by the studies that are available. It is worth noting that study authors in this field face ethical challenges when seeking to carry out research with young people who are in the midst of an eating disorder.

One additional limitation relates to time; more recent primary studies may now have been published, which appears to be a rapidly growing area of research [anonymity no author]. Digital platforms, distinct from those discussed in the research identified to date, may now be preferred by users more. The review also draws attention to the need for further engagement with this complexity, given young people’s ongoing relationship with digital online platforms and the subjectivity of their experiences in those spaces.

The synthesis highlighted the gender imbalance in the studies, with only one study out of 13 providing a young man’s perspective. This suggests that there are evidential gaps here and that further investigation into male experiences of EDs and body norms is needed. There was also no demographic data reported on sexuality or exploration of the experiences of the LGBTQIA+ community, despite body image being a significant issue for this community. Although there were five studies with participants from different ethnic groups, the number of participants taking part was small. It was also difficult to discern from the data the ethnicity of individuals and whether that this had any bearing on their lived experience. This lack of disclosure could be to protect the individual. However, it would be useful for future research to collect data from ethnic minority participants and members of the LGBTQIA+ community purposively, to explore whether their cultural/religious upbringings or identities play any role in the types of ED content they engage with and the nature of this engagement.

As we see an ever-increasing number of younger children using digital devices and gaining access to online platforms, it seems increasingly important to investigate this within the ED context. The more recent literature made steps in including young people aged 17 and under. This is an extremely sensitive topic and considerable attention to ethical care is needed when exploring lived experience of eating disorders, especially among younger young people. The ‘Community’ theme could also be examined further within the context of preventative intervention research, which some participants alluded to. Finally, in terms of media platforms, the studies in the review primarily analysed data about Instagram, with only one study each exploring TikTok and Snapchat. However new platforms are emerging all the time and there is a need for both researchers and reviewers to somehow keep abreast of the platforms young people are actually using.

## Conclusion

This is the first qualitative evidence synthesis to examine online eating disorder content and provides an important account of how young people experience online eating disorder and idealised body image content. The findings highlight synergies in engagement around the key themes such as ‘community’ which had not yet been explored to any depth within the context of online eating disorder content. This paper also draws attention to the need for further engagement with this complexity, given the ongoing relationship young people have with digital online platforms and the subjectivity of their experiences in those spaces. New and existing policy interventions focused on supporting young people in this area could consider whether professionals routinely explore the role of online platforms as a tool that can support or hinder recovery from eating disorders. There is much potential for initiatives where those with lived experience of eating disorders and the use of online content work with health professionals and researchers to co-design online spaces and campaigns (58). Digital literacy interventions, particularly when related to online harms, could support young people to understand not only that engagement with certain types of content can be detrimental to their mental health, including body image and self-esteem, but also that identifying the line between helpful and unhelpful is subjective. Therefore, it can be difficult to determine and could require outside help to decipher (for example, by consulting with family, friends, or teachers). Given that young people often go online to seek information and support, there is not only scope for online platforms, such as social media and networking sites, to direct young people towards professional help, but also for platforms, including mobile applications, to become tools for increasing the accessibility of help for young people. The design of digital interventions could also be informed by how young people use online spaces, to develop a person-centred and strength-based approach to addressing eating disorders and body image concern. Furthermore, the awareness demonstrated by young people about online content and inauthenticity, competitiveness and self-critiquing requires further scope for alternative-evidence based prevention/intervention approaches (for example, self-compassion, Goss & Allan 2014).

## Acknowledgements

We would like to thank the advisory panel members and the young people who took part in our consultation workshops for their valuable contributions. We would also like to thank the organisations who helped recruit young people (Fixers UK, Hearts and Minds, and The McPin Foundation) and Dr Louca-Mai Brady for her work helping devise and facilitate the workshops. Finally, we would like to thank the research assistants Jia Han, Rafael Rivero Labrador, Jonathan Allen and Juliette O’Connell for their contribution to the earlier stages of the review.

## CRediT authorship contribution statement

All authors contributed to the conceptualisation and design of the review and writing of the manuscript. KD developed and wrote the initial draft of the protocol. CS designed and executed the search strategy and screened some titles and abstracts. MK, RR, and KD, screened titles and abstracts, retrieved and screened full texts and descriptively coded studies. MK and RR performed data extraction and quality assessment of the included studies. MK and RR planned and conducted the write-up of the synthesis. MK led the writing of the discussion and conclusion, with contributions from RR and KD. MK prepared the manuscript for submission. All authors reviewed the manuscript and gave final approval for submission.

## Supporting information

Figure 1: Flow of studies

Supplementary Files A Included Studies

Supplementary Files B:Details of Included Studies

## Declaration of Competing Interest

None.

## Funding

This paper is funded through the NIHR PRP contract with the EPPI Centre at UCL (Evidence review facility to support national policy development and implementation, NIHR200701). The views expressed are those of the author(s) and not necessarily those of the NIHR or the Department of Health and Social Care.

## Data Availability

Data will be made available on request.

## Notes

### Competing Interest Statement

The authors have declared no competing interest.

### Funding Statement

Yes

### Author Declarations

Calum Gordon On behalf of UCL Institute of Education Research Ethics Committee Research Ethics Officer UCL Institute of Education

